# Bayesian genome-wide TWAS with reference transcriptomic data of brain and blood tissues identified 93 risk genes for Alzheimer’s disease dementia

**DOI:** 10.1101/2023.07.06.23292336

**Authors:** Shuyi Guo, Jingjing Yang

## Abstract

**Background:** Transcriptome-wide association study (TWAS) is an influential tool for identifying novel genes associated with complex diseases, where their genetic effects may be mediated through transcriptome. TWAS utilizes reference genetic and transcriptomic data to estimate genetic effect sizes on expression quantitative traits of target genes (i.e., effect sizes of a broad sense of expression quantitative trait loci, eQTL). These estimated effect sizes are then employed as variant weights in burden gene-based association test statistics, facilitating the mapping of risk genes for complex diseases with genome-wide association study (GWAS) data. However, most existing TWAS of Alzheimer’s disease (AD) dementia have primarily focused on *cis*-eQTL, disregarding potential *trans*-eQTL. To overcome this limitation, we applied the Bayesian Genome-wide TWAS (BGW-TWAS) method which incorporated both *cis*- and *trans*-eQTL of brain and blood tissues to enhance mapping risk genes for AD dementia.

**Methods:** We first applied BGW-TWAS to the Genotype-Tissue Expression (GTEx) V8 dataset to estimate *cis*- and *trans*-eQTL effect sizes of the prefrontal cortex, cortex, and whole blood tissues. Subsequently, estimated eQTL effect sizes were integrated with the summary data of the most recent GWAS of AD dementia to obtain BGW-TWAS (i.e., gene-based association test) p-values of AD dementia per tissue type. Finally, we used the aggregated Cauchy association test to combine TWAS p-values across three tissues to obtain omnibus TWAS p-values per gene.

**Results:** We identified 37 genes in prefrontal cortex, 55 in cortex, and 51 in whole blood that were significantly associated with AD dementia. By combining BGW-TWAS p-values across these three tissues, we obtained 93 significant risk genes including 29 genes primarily due to *trans*-eQTL and 50 novel genes. Utilizing protein-protein interaction network and phenotype enrichment analyses with these 93 significant risk genes, we detected 5 functional clusters comprised of both known and novel AD risk genes and 7 enriched phenotypes.

**Conclusion:** We applied BGW-TWAS and aggregated Cauchy test methods to integrate both *cis*- and *trans*-eQTL data of brain and blood tissues with GWAS summary data to identify risk genes of AD dementia. The risk genes we identified provide novel insights into the underlying biological pathways implicated in AD dementia.

## Background

Alzheimer’s disease (AD) dementia is a complex neurodegenerative disorder characterized by progressive cognitive decline and memory loss, currently affecting 6.5 million Americans aged 65 and older. AD dementia is listed as the seventh-leading cause of death in the United States of America [1]. Despite extensive research, the underlying biological mechanisms of AD dementia remain elusive, and effective treatments are still lacking [2]. Recent studies have highlighted the important roles of genomic risk factors of AD dementia [3, 4]. The most recent genome-wide association study (GWAS) identified a total of 38 risk genes for AD dementia [5]. However, these identified GWAS risk genes explain only a small portion of the heritability of AD dementia, suggesting more risk genes might contribute to disease risk. Also, the biological mechanisms underlying the majority of GWAS risk genes remain unknown. Transcriptome-wide association studies (TWAS) have emerged as an influential tool for identifying risk genes associated with complex diseases, particularly those with genetic effects mediated through transcriptome [6, 7]. For example, a recent TWAS of AD dementia by Sun et al. has identified 53 risk genes by standard two-stage TWAS methods [8].

Standard two-stage TWAS methods first train gene expression prediction regression models by using reference genetic and transcriptomic data profiled of the same training samples, taking quantitative gene expression traits as response variables and genetic variants as predictors (Stage I). Estimated effect sizes of genetic variants in the gene expression prediction models could be viewed as effect sizes of a broad sense of expression quantitative trait loci (eQTL), which will then be taken as variant weights in burden gene-based association test statistics to map risk genes with GWAS data (Stage II). The TWAS association test in Stage II is equivalent to testing the association between predicted genetically regulated gene expression (GReX) levels from GWAS data and the phenotype of interest in the test cohort (i.e., GWAS data) [9].

However, one limitation of most existing TWAS methods is that they only consider *cis*-eQTL [6, 10, 11], genetic variants located within the 1MB region around the target gene, in the gene expression prediction models. The limitation is mainly due to the computational bottleneck of considering genome-wide genetic variants to fit gene expression prediction models for transcriptome-wide ∼20K genes per tissue type. The resulting cavity is failing to account for *trans*-eQTLs located outside of the 1MB region around the target gene that have been found to play important roles in biological processes and disease susceptibility [12, 13]. Incorporating *trans*-eQTLs in TWAS is essential as they can reveal additional regulatory mechanisms, help identify novel associations between GReX and the phenotype of interest, and further our understanding of the underlying biological mechanisms of complex diseases.

To overcome this limitation for studying AD dementia, we employed the Bayesian Genome-wide TWAS (BGW-TWAS) method that incorporated both *cis*- and *trans*-eQTL for TWAS [14]. We applied BGW-TWAS to the reference Genotype-Tissue Expression (GTEx) V8 dataset [15] of three tissues –– prefrontal cortex, cortex, and whole blood. The selection of prefrontal cortex and cortex tissues was based on substantial evidence linking their involvement to the progression of AD dementia [16]. The selection of whole blood tissue was due to three reasons: i) a large sample size exists (n=574) for the whole blood tissue in the reference GTEx V8 dataset; ii) recent studies have demonstrated that specific gene expression products in whole blood can serve as biomarkers for AD dementia [17, 18]; iii) gene expression in the whole blood and that of the brain’s cortex were found correlated [19].

Specifically, *cis*- and *trans*-eQTL effect sizes of prefrontal cortex, cortex, and whole blood tissues were first estimated by BGW-TWAS. Second, estimated eQTL effect sizes were integrated with the summary data of the most recent GWAS of AD dementia (n=∼762K, excluding 23&me samples) [5] to calculate BGW-TWAS (i.e., gene-based association test) p-values of AD dementia per tissue type, using the S-PrediXcan test statistic [20]. Third, we used the omnibus aggregated Cauchy association test (ACAT-O) [21] to combine TWAS p-values across three tissues to obtain omnibus TWAS p-values per gene. The workflow is presented in **Figure 1**.

**Figure 1.**
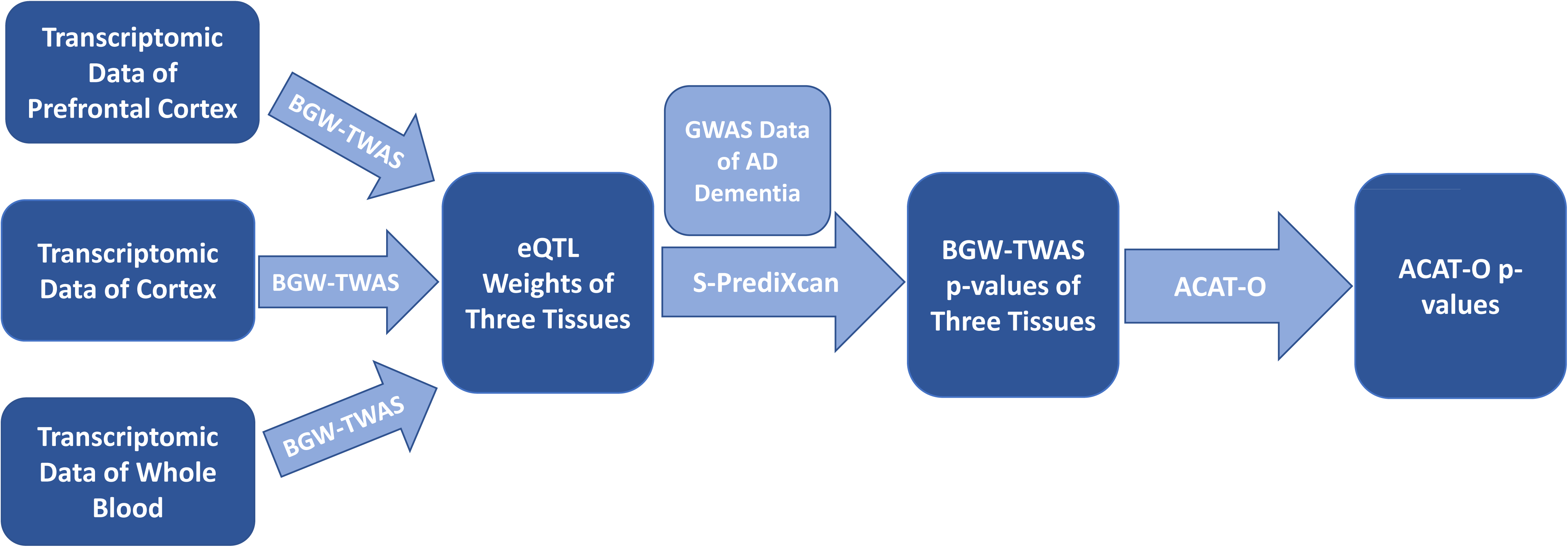
Study workflow.

As a result, we identified 37 significant genes with the prefrontal cortex, 55 significant genes with the cortex, and 51 significant genes with the whole blood reference transcriptomic data. Combining BGW-TWAS p-values across these three tissues by ACAT-O, we obtained a total of 93 significant TWAS risk genes for AD dementia, including several well-known AD risk genes and 50 novel genes not detected by previous GWAS nor TWAS. Through protein-protein association network analysis [22] with these 93 significant TWAS risk genes, we detected a main functional cluster comprised of known AD risk genes including *APOE*, *BIN1*, *CASS4*, *MS4A4A*, *MS4A6A*, *SLC24A4*, *CD33*, *HLA-DRB1* and our novel findings. We also identified another 4 clusters comprising both known and novel risk genes of AD. In addition, these 93 significant TWAS risk genes were found enriched with known risk genes of 7 phenotypes, including Apolipoprotein B, low-density lipoprotein cholesterol, inflammatory biomarkers, and C-reactive protein.

## Methods

### Bayesian Genome-wide TWAS (BGW-TWAS)

BGW-TWAS [14] is a recently proposed TWAS method that incorporates both *cis*- and *trans*-genetic variants of the target genes as predictors in the gene expression prediction models (Stage I) as follows:

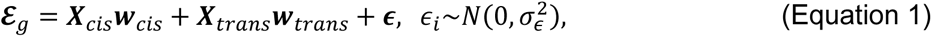

where *ε_g_* denotes the target gene expression trait, (***X**_cis_*, ***X**_trans_*) denotes the genotype data matrix of *cis*- and *trans*-genetic variants; (***w**_cis_*, ***w**_trans_*) denotes the corresponding *cis*- and *trans*-effect sizes; and *ϵ* denotes the errors following a normal distribution with mean 0 and variance 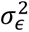. Here, we briefly describe the BGW-TWAS method.

BGW-TWAS assumes the Bayesian variable selection regression (BVSR) model [23] to enforce sparse eQTL models, by assuming specific spike-and-slab prior distributions for *cis*- and *trans*-effect sizes given by

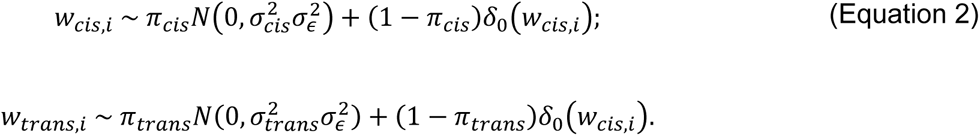

Here, (*π_cis_*, *π_trans_*) denote the respective probability that the corresponding effect size is normally distributed, and *δ*_0_(*w_i_*) is the point mass density function that takes value 0 when *w_i_* ≠ 0 and 1 when *w_i_* = 0.

BGW-TWAS employs multiple computational strategies to enable efficient computation to account for genome-wide genetic variants as predictors in the gene expression prediction models. A previously developed scalable expectation-maximization Markov Chain Monte Carlo (EM-MCMC) algorithm [24] is adapted and used by BGW-TWAS. Genome-wide variants are first segmented into approximately independent genome blocks. Genome blocks containing *cis*-genetic variants or containing top *trans*-eQTL with single variant test p-values < 10^−5^ will be selected for implementing the EM-MCMC algorithm to estimate eQTL effect sizes in the joint multivariable regression model as in Equation 1. The posterior causal probabilities (PCP) for “eQTL” with non-zero posterior effect size estimates will also be estimated. The product of estimated PCPs and effect sizes will represent the expected posterior effect sizes and be used as variant weights in the follow-up TWAS tests (Stage II). The details of the BGW-TWAS method are referred to the BGW-TWAS paper [14].

### Gene-based Association Test by S-PrediXcan Test Statistic

With *cis*- and *trans*-eQTL weights estimated by BGW-TWAS method and summary-level GWAS data (i.e., Z-scores by single variant tests), we employed the S-PrediXcan [20] approach to calculate the burden type TWAS Z-score test statistic *Z_g_* per gene as follows:

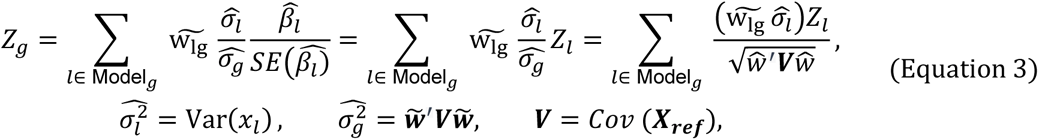

where 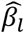 denotes the effect size of genetic variant *l* from GWAS, *Z_l_* denote the Z-score statistic by single variant GWAS test, and 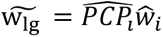 is the weight of genetic variant *l* estimated by BGW-TWAS. Here, ***X**_ref_* denotes the genotype matrix from a reference panel, and ***V*** is the genotype covariance matrix. Two-tailed BGW-TWAS p-values can then be obtained from the TWAS Z-score test statistics as in Equation 3.

### Omnibus Aggregated Cauchy Association Test (ACAT-O)

ACAT-O is an omnibus test that combines p-values of multiple tests for the same hypothesis [21], which employs a linear combination of transformed p-values as the test statistic. As shown in the following formula,

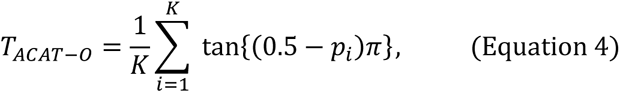

where *p_i_*s are the p-values of K tests and the ACAT-O statistic *T_ACAT-0_* follows a standard Cauchy distribution under the null hypothesis. As shown in **Figure 1**, in this study, ACAT-O method was used to combine BGW-TWAS p-values per gene across three considered tissues to obtain combined TWAS p-values. Combined TWAS p-values were obtained by using the “ACATO” function from the R package “sumFREGAT” [25].

### Genotype-Tissue Expression (GTEx) Data

The publicly available GTEx V8 (dbGaP phs000424.v8.p2) data [15] contain whole genome sequencing (WGS) data of 838 human donors and RNA sequencing transcriptomic data of 17,382 normal samples from 52 human tissues and two cell lines. We used the transcriptomic data of the prefrontal cortex (n=158), cortex (n=184), and whole blood (n=574) tissues and the corresponding WGS data as reference panel to estimate *cis*- and *trans*-eQTL weights by BGW-TWAS. In particular, genetic variants with missing rate < 20%, minor allele frequency > 0.01, and Hardy-Weinberg equilibrium p-value > 10^−5^ were considered for fitting the gene expression prediction models. Gene expression data of Transcripts Per Million (TPM) per sample per tissue were downloaded from the GTEx portal. Genes with > 0.1 TPM in ≥ 10 samples were considered. Raw gene expression data (TPM) were log 2 transformed and then adjusted for age, body mass index (BMI), top five genotype principal components, and top probabilistic estimation of expression residuals (PEER) factors [9]. Adjusted gene expression quantitative traits were then taken as response variables in the gene expression prediction model.

### Summary-level GWAS Data of AD Dementia

The summary-level GWAS data of AD dementia (i.e., single variant Z-score test statistics obtained by meta-analysis) were generated by the latest GWAS by Wightman et al [5], including ∼ 762K individuals from 12 cohorts except for 23&Me samples. About 11.3% of participants had clinically diagnosed AD dementia.

### Protein-Protein Interaction Network and Pathway Analysis

STRING (version 11.5) [22] is a bioinformatics web tool that provides information on protein-protein interactions (PPI) and networks, as well as functional characterization of genes and proteins. The tool integrates different types of evidence from public databases, such as genomic context, high-throughput experiments, and previous knowledge from other databases, to generate reliable predictions of protein interactions and build networks and pathways. Provided with a list of gene names of our significant TWAS risk genes, STRING will construct networks based on the protein-protein interactions of the corresponding proteins, as well as identify phenotypes that have risk genes enriched in the provided list. Proteins corresponding to provided genes are considered as nodes in the PPI network. Protein-protein edges represent the predicted functional associations, colored differently to indicate seven categories –– co-expression, text-mining, experiments (biochemical/genetic data), databases (previously curated pathway and protein complex information), gene co-occurrence, gene fusion and gene neighborhood. Gene co-occurrence, fusion, and neighborhood represent association predictions are based on whole-genome comparisons [26].

## Results

### BGW-TWAS Results of AD Dementia

Using BGW-TWAS method, we trained gene expression prediction models for 23,721 genes of prefrontal cortex tissue, 23,864 genes of cortex tissue, and 19,514 genes of whole blood tissue. We identified 37, 55, and 51 TWAS risk genes with significant p-values (with Bonferroni correction) for prefrontal cortex, cortex, and whole blood tissues, respectively. Of these, 15 genes were significant for prefrontal cortex and cortex, 6 were significant for prefrontal cortex and whole blood, 8 were significant for cortex and whole blood, and 3 were significant for all three tissue types (Supplemental Figure 1). Manhattan plots of BGW-TWAS results of the three tissues were presented in Supplemental Figures 2-4.

We also summarized the proportion of *trans*-eQTL with non-zero weights for all significant genes of three tissues in Supplemental Figure 5. There were 12 (32.4%), 24 (43.6%), and 16 (31.4%) significant genes with >50% *trans*-eQTLs in prefrontal cortex, cortex, and whole blood tissues, respectively. These results demonstrated that *trans*-eQTL had important contributions to significant TWAS risk genes of all three tissues.

### ACAT-O Results of AD Dementia

Combining BGW-TWAS p-values across three tissues per gene by ACAT-O, we obtained ACAT-O p-values for a total of 17,468 genes. We identified 93 genes with significant ACAT-O p-values (with Bonferroni correction). As illustrated by the Manhattan plot (**Figure 2**), 43 genes were located on chromosome 19 around the well-known *APOE* locus, which is consistent with prior TWAS findings [8, 27]. Additionally, we observed clusters of significant genes on chromosomes 6, 7, and 11.

**Figure 2.**
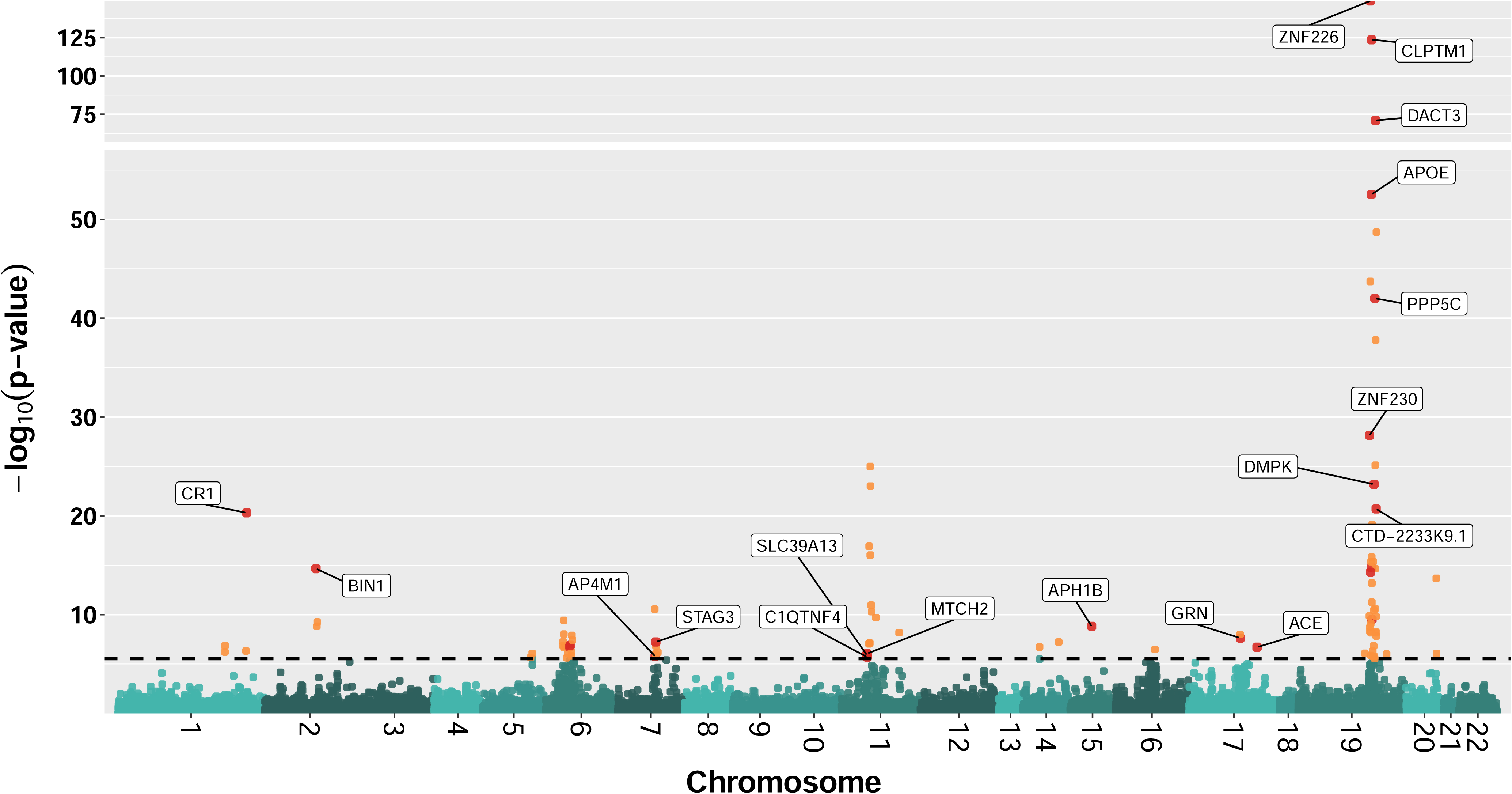
Manhattan plot of ACAT-O TWAS p-values for studying AD dementia. The horizontal dashed line represents the Bonferroni corrected significance threshold of the p-value, given by dividing 0.05 by the total number of test genes. Orange dots indicate genes that were significant in only one tissue type, while red dots with labels highlight genes that were significant in at least two tissue types.

To compare with TWAS results using only *cis*-eQTL, we conducted TWAS for all three tissue types using only *cis*-eQTL with non-zero weights estimated by BGW-TWAS, and then combined these TWAS p-values per gene by ACAT-O. We found that 64 of the 93 significant genes (using both *cis*- and *trans*-eQTL) retained their significance using only *cis*-eQTL, while 29 genes were no longer significant (Supplemental Figure 6). These results indicated that ∼31% of significant TWAS genes were primarily driven by *trans*-eQTL. We presented example significant TWAS risk genes (58 out of all 93) in **Table 1**, categorizing them based on whether their significances were primarily driven by *cis*-eQTL or *trans*-eQTL. The remaining TWAS risk genes were presented in Supplemental Table 1.

**Table 1.** Example 58 TWAS risk genes of AD dementia with significant ACAT-O p-values, which were mainly driven by *cis*-eQTL or *trans*-eQTL. This table includes all the significant genes primarily driven by *trans*-eQTL (29 genes in the right column) and part of the significant genes driven by *cis*-eQTL (29 genes in the left column).

| Significance Driven by *cis*-eQTL |  |  | Significance Driven by *trans*-eQTL |  |  |
| --- | --- | --- | --- | --- | --- |
| Gene | CHR | ACACT-O p-value | Gene | CHR | ACAT-O p-value |
| CR1<sup>ab</sup> | 1 | 4.99E-21 | RP11-343H5.4<sup>b</sup> | 1 | 4.80E-07 |
| BIN1<sup>bc</sup> | 2 | 2.22E-15 | DDR1<sup>a</sup> | 6 | 2.08E-07 |
| HLA-DRB1<sup>ab</sup> | 6 | 1.74E-07 | BAG6<sup>c</sup> | 6 | 1.69E-07 |
| TAP2<sup>ac</sup> | 6 | 1.28E-07 | ATAT1<sup>b</sup> | 6 | 1.43E-07 |
| AP4M1<sup>ab</sup> | 7 | 1.67E-06 | DHX16<sup>a</sup> | 6 | 5.46E-08 |
| STAG3<sup>ab</sup> | 7 | 5.91E-08 | XXbac-BPG252P9.10<sup>a</sup> | 6 | 9.65E-09 |
| C1QTNF4<sup>bc</sup> | 11 | 2.00E-06 | TUBB<sup>b</sup> | 6 | 3.88E-10 |
| SLC39A13<sup>ab</sup> | 11 | 1.19E-06 | CTNND1<sup>a</sup> | 11 | 9.02E-08 |
| MTCH2<sup>ab</sup> | 11 | 8.79E-07 | SLC3A2<sup>c</sup> | 11 | 2.01E-10 |
| MS4A3<sup>c</sup> | 11 | 9.82E-17 | VWCE<sup>b</sup> | 11 | 4.95E-11 |
| MS4A4A<sup>c</sup> | 11 | 9.99E-24 | AP001350.4<sup>b</sup> | 11 | 1.22E-17 |
| MS4A6A<sup>c</sup> | 11 | 1.03E-25 | RP5-882C2.2<sup>a</sup> | 17 | 1.03E-08 |
| APH1B<sup>ac</sup> | 15 | 1.56E-09 | PGLYRP1<sup>a</sup> | 19 | 2.82E-06 |
| ACE<sup>abc</sup> | 17 | 2.00E-07 | CTC-435M10.12<sup>c</sup> | 19 | 8.44E-07 |
| GRN<sup>ab</sup> | 17 | 2.37E-08 | SNRPD2<sup>b</sup> | 19 | 1.52E-07 |
| GEMIN7<sup>ab</sup> | 19 | 3.09E-10 | ARHGAP35<sup>c</sup> | 19 | 1.45E-08 |
| CTB-179K24.3<sup>c</sup> | 19 | 6.47E-14 | TMEM160<sup>c</sup> | 19 | 6.75E-09 |
| ZNF285<sup>bc</sup> | 19 | 5.11E-15 | CTB-12A17.2<sup>a</sup> | 19 | 5.35E-09 |
| APOC2<sup>abc</sup> | 19 | 1.78E-15 | SLC1A5<sup>b</sup> | 19 | 1.49E-10 |
| BCAM<sup>c</sup> | 19 | 4.44E-16 | AC007191.4<sup>b</sup> | 19 | 3.59E-11 |
| GPR4<sup>a</sup> | 19 | 4.44E-16 | CALM3<sup>b</sup> | 19 | 2.44E-11 |
| RELB<sup>c</sup> | 19 | 1.51E-16 | GNG8<sup>b</sup> | 19 | 2.22E-15 |
| DMPK<sup>ab</sup> | 19 | 6.52E-24 | TRAPPC6A<sup>c</sup> | 19 | 8.13E-20 |
| ZNF230<sup>bc</sup> | 19 | 7.13E-29 | CTD-2233K9.1<sup>ab</sup> | 19 | 2.00E-21 |
| CTC-204F22.1<sup>b</sup> | 19 | 1.91E-44 | PTGIR<sup>c</sup> | 19 | 7.70E-26 |
| APOE<sup>bc</sup> | 19 | 3.03E-53 | DACT3-AS1<sup>a</sup> | 19 | 1.61E-38 |
| CLPTM1<sup>abc</sup> | 19 | 2.29E-124 | PPP5C<sup>ab</sup> | 19 | 9.86E-43 |
| ZNF226<sup>ac</sup> | 19 | 1.14E-149 | ZC3H4<sup>a</sup> | 19 | 2.04E-49 |
| CASS4<sup>c</sup> | 20 | 2.15E-14 | DACT3<sup>ab</sup> | 19 | 1.19E-71 |

a: Genes significant for prefrontal cortex tissue
b: Genes significant for cortex tissue
c: Genes significant for whole blood tissue

### Contributing eQTL of Significant TWAS Risk Genes

To investigate how eQTL contributed to significant TWAS risk genes, we plotted eQTL weights estimated by BGW-TWAS for three tissues of several example TWAS risk genes in **Figure 3**. Specifically, column A in **Figure 3** shows the eQTL weights (in three tissues) of gene *ACE* whose significance is primarily due to *cis*-eQTL; column B shows the eQTL weights (in three tissues) of gene *DACT3* whose significance is primarily due to *trans*-eQTL; and column C shows the eQTL weights of three genes (*CTNND1*, *AP001350.4*, and *SLC3A2*) in prefrontal cortex, cortex, and whole blood tissues, respectively. As shown in **Figure 3**, each dot represents one eQTL with colors ranging from yellow to red to represent the corresponding AD dementia GWAS p-value. We can see that *cis*- or *trans*-eQTL colocalizing with potential significant GWAS p-values (colored from yellow to red) are driving the significant TWAS association of the test gene. It is noticeable that all the *trans*-eQTL of the example genes (and actually for most of the genes we studied) are still on the same chromosome as the test gene.

**Figure 3.**
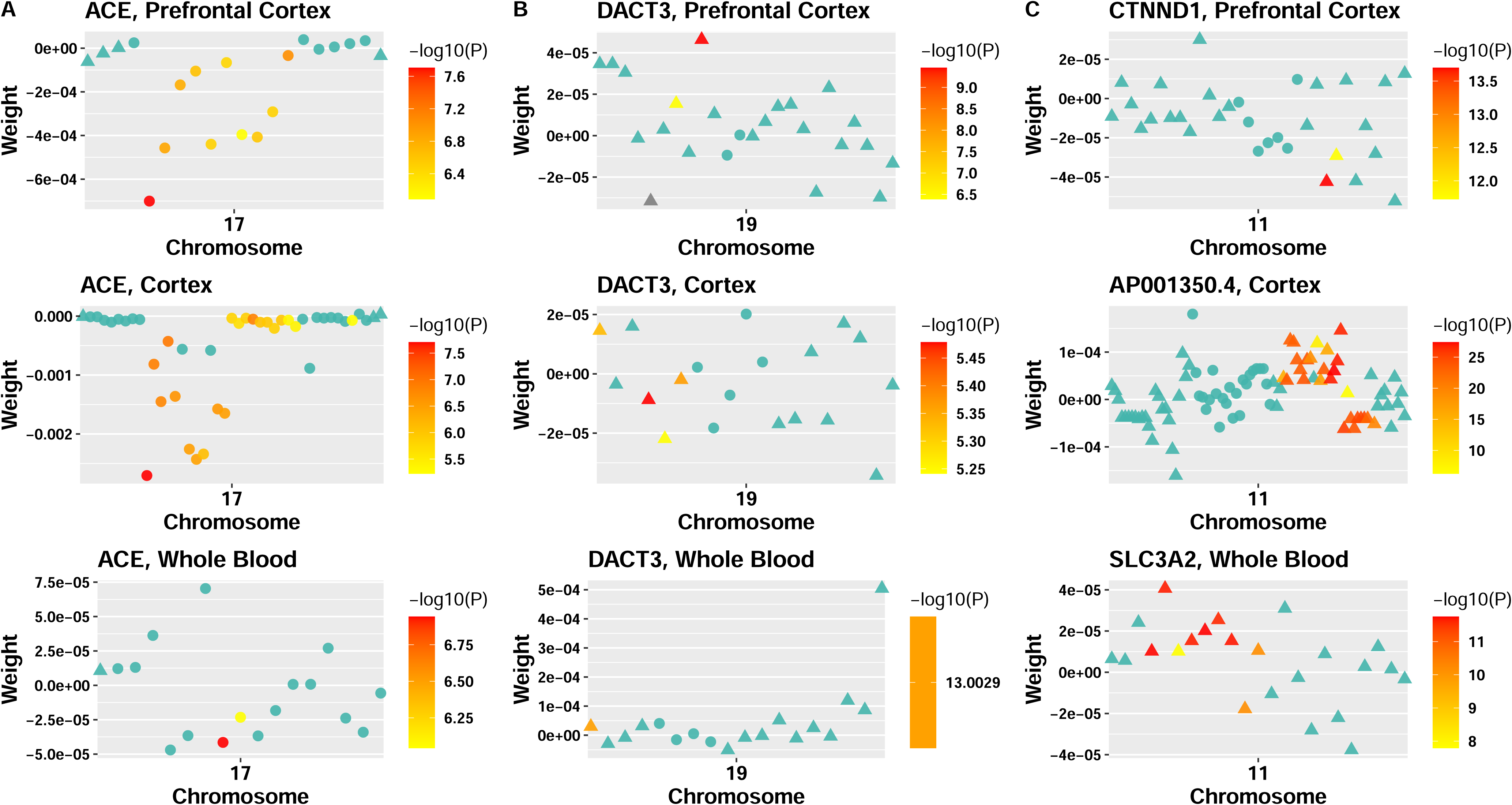
Scatter plots of eQTL weights estimated by BGW-TWAS of example TWAS risk genes for three tissues. Column A: gene *ACE* in three tissues; Column B: gene *DACT3* in three tissues; Column C: gene *CTNND1*, *AP001350.4* and *SLC3A2*, in prefrontal cortex, cortex, and whole blood tissues, respectively. Y-axis depicts the values of eQTL weights estimated by BGW-TWAS, and the x-axis shows the base pair position of the corresponding eQTL. Solid circles denote *cis*-eQTL, and triangles refer to *trans*-eQTL. Color legend denotes the -log (GWAS p-value) of the corresponding eQTL.

**Figure 4.**
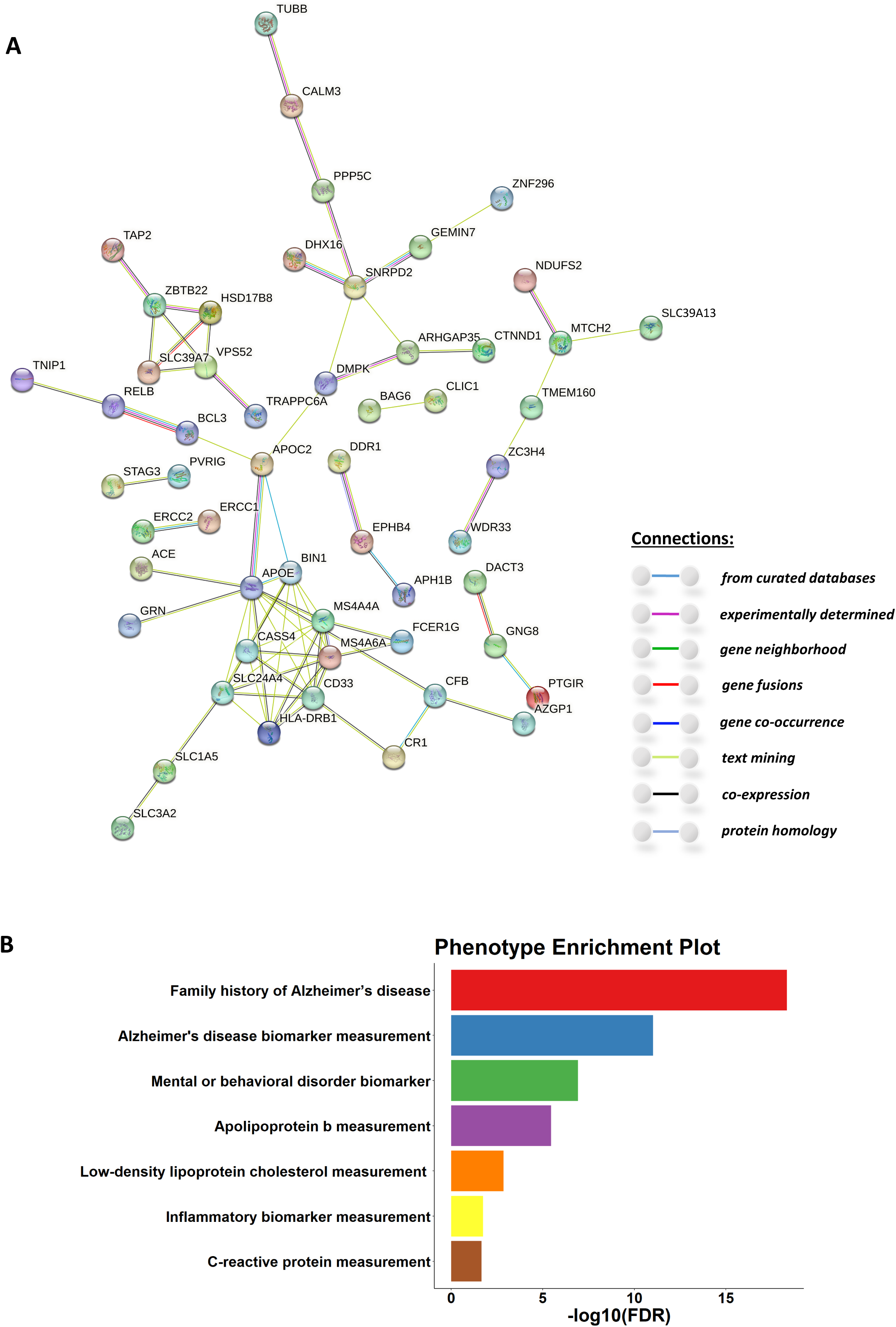
Protein-protein interaction networks of all 93 significant TWAS risk genes and phenotype enrichment analysis results. A: protein-protein interaction network of the 93 significant TWAS risk genes generated by STRING, with the edge color legend shown in the right bottom; B: -log10 of the false discovery rate (FDR, x-axis) for testing the enrichment of known risk genes of the corresponding phenotype (y-axis) in the list of 93 significant TWAS risk genes.

### Known Risk Genes for AD Dementia

Next, we compared our TWAS findings using both *cis*- and *trans*-eQTL and three tissues to the TWAS findings by TIGAR [9, 11] using only *cis*-eQTL, prefrontal cortex tissue [28], and the same GWAS summary data of AD dementia [26]. We found that 9 out of these 93 TWAS risk genes detected in this study were identified by both TWAS approaches, including *HLA-DRB1* on chromosome 6, *OSBP* on chromosome 11, *ACE* on chromosome 17, as well as *BCL3*, *CLPTM1*, *DMPK*, *GEMIN7*, *ZNF230*, and *ZNF296* on chromosome 19. Additionally, 34 out of the 93 TWAS risk genes detected in this study were also reported by previous TWAS or GWAS studies as being related to AD [4, 5, 8, 29, 30], such as known risk genes of *ACE*, *APOC2*, and *CLPTM1* that were found significant by BGW-TWAS with all three tissue types.

Interestingly, among well-known genes related to AD dementia that were also identified as significant TWAS risk genes in this study, we observed that the GReX of *APOE* in cortex tissue was positively associated with risk of AD dementia, while having a significant negative association in whole blood. The GReX of *APOC2* on chromosome 9 was found to be positively associated with risk of AD dementia in prefrontal cortex and cortex, while having a significant negative association in whole blood. The GReX of *ACE* and *CLPTM1* was positively associated with risk of AD dementia in all three tissues.

In addition to well-known risk genes related to AD dementia, we also identified genes that have been detected in recent TWAS of AD. For instance, a TWAS conducted with hippocampus tissue [30] showed that the expression of *DACT3*, *SNRPD2,* and *DMPK* in the hippocampus affected the risk of AD, which were also identified as significant TWAS risk genes in this study. Moreover, a recent study that integrated eQTL data of blood tissue and GWAS of late-onset AD (LOAD) by a Bayesian statistical method [31] identified risk gene *ZNF226*, which was also detected in this study. Gene *TNIP1* with significant BGW-TWAS p-value in whole blood has recently been identified as containing significant eQTL of AD within blood tissue [5].

### Novel TWAS Risk Genes of AD Dementia

We found that 50 out of these 93 significant TWAS risk genes of AD dementia were novel findings, including 26 genes whose significance were due to *trans*-eQTLs that have not been accounted for by prior TWAS. Previous biological studies provided insights of some of our novel findings with respect to AD dementia. For example, the protein expression of *UGGT1,* a gene coding N-glycosylation-related proteins in the endoplasmic reticulum, was found upregulated in AD brain capillaries [32]. Gene *BAG6* was found to prevent the accumulation of neurodegeneration-associated fragments of TAR DNA-binding protein 43 (TDP43) which is one type of AD pathology [33]. Gene *DHX16* was also recognized as a candidate risk gene for AD [34, 35], and the RNA helicase Dhx16 induces transcription alterations and DNA methylation changes that are involved in memory-related neurological and neuropsychiatric disorders [36]. Also, gene *TUBB* encodes a β-tubulin protein that forms a dimer with alpha tubulin and acts as a structural component of microtubules, where β-tubulin was found aggregating in AD cases [37].

### Protein-protein Interaction Network Analysis by STRING

To further understand the underlying biological mechanisms of our identified 93 TWAS risk genes, we constructed protein-protein interaction (PPI) networks and performed phenotype enrichment analysis using the STRING tool. With our identified 93 TWAS risk genes, a total of 5 functional clusters were identified by STRING (**Figure 4A**), including a major functional cluster composed of known AD risk genes *APOE*, *BIN1*, *CASS4*, *MS4A4A*, *MS4A6A*, *SLC24A4*, *CD33*, and *HLA-DRB1*. Besides these known AD risk genes, several of our novel TWAS risk genes were found connected within the main cluster. For example, novel genes *SLC1A5* and *SLC3A2* were connected to *SLC24A34*; novel genes *CFB* and *AZGP1* were connected to *APOE*. Three pathways were found to be enriched with TWAS risk genes in the main cluster, including the CD20-like family/membrane-spanning 4-domains (MS4) subfamily and MHC class I protein-binding genes that are involved with immune system processes. These two immune system related pathways include B-cell surface antigen CD20/MS4A and MHC class I proteins that provide antigens to T cells. It is noticeable that known AD risk genes *MS4A4A* and *MS4A6A* are among the CD20-like family/MS4A pathway [38], and MHC class I proteins are found to be critical in the context of aging brains and AD [39, 40]. The third pathway involves simultanagnosia and sodium/potassium/calcium exchanger 4, including genes *SLC24A4, CD33*, *CASS4*, and *MS4A4A*. Previous studies showed that AD risk variants in *SLC24A4* were correlated with increased expression in blood and brain regions [41]. Our study also found that the GReX of *SLC24A4* in the whole blood was positively correlated with risk of AD dementia.

In addition to the main cluster discussed above, two other clusters are associated with the known AD risk gene *APOC2*. One cluster includes known AD risk genes *BCL3*, *RELB*, and *TNIP1*, and the second one includes *DMPK*, *ARHGAP35*, *CTNND1*, *SNRPD2*, *DHX16*, *GEMIN7*, *ZNF296*, *PPP5C*, *CALM3*, and *TUBB*. Both *BCL3* and *RELB* have known AD risk variants near the *APOE* region [42], and AD-related eQTL have been detected within *TNIP1* [5]. Among the genes in the second cluster, *DMPK*, *SNRPD2, GEMIN7,* and *ZNF296* are known AD risk genes, with the remaining being novel findings. In particular, *DMPK* and *SNRPD2* have been identified as AD risk genes in the hippocampus and putamen by a previous TWAS [30]. Genes *GEMIN7* and *ZNF296* have been identified as AD risk genes in a TWAS conducted on the cortex and the amygdala region of the brain [8], respectively.

Another PPI network worth paying attention to is the cluster composed of *TAP2*, *ZBTB22*, *HSD17B8*, *SLC39A7*, *VPS52,* and *TRAPPC6A*. Except for *TRAPPC6A* [43], other genes are novel findings located on chromosome 6. Interestingly, gene *ZBTB22* encodes a transcription factor containing both zinc finger and *BTB* domains, and gene *SLC39A7* encodes a zinc transporter protein. Previous studies showed that some zinc finger proteins influenced the accumulation of neurofibrillary tangles, one type of AD pathology [44]. Gene *TAP2* acts as a molecular scaffold for the final stage of MHC class I folding which was detected as a potential AD-related function [40].

Another cluster is composed of known AD risk genes *MTCH2* and *NDUFS2,* as well as novel findings *WDR33*, *ZC3H4*, *TMEM160*, and *SLC39A13*. In particular, a recent study has shown that decrease of *MTCH2* expression level in the forebrain (playing a crucial role in mitochondrial metabolism) could impair cognitive functions related to the hippocampus and might eventually lead to AD [45]. Similarly, gene *NDUFS2* has been shown related to the oxidative phosphorylation process of mitochondrial metabolism which affects the risk of AD [17].

### Phenotype Enrichment Analysis

The phenotype enrichment analysis by STRING (**Figure 4B**) found that our identified 93 TWAS risk genes were enriched with known risk genes of family history of AD (false discovery rate, FDR = 4.57e-19), mental or behavioral disorders (FDR = 1.18e-07), AD biomarkers (FDR = 9.52e-12), Apolipoprotein b (ApoB, FDR = 3.44e-06), low-density lipoprotein cholesterol (LDL-C, FDR = 1.40e-03), inflammatory biomarkers (FDR = 1.79e-02), and C-reactive protein (CRP, FDR = 2.16e-02). For example, TWAS risk genes *APOE*, *BIN1*, and *CR1* are known risk genes for family history of AD; *HLA-DRB1*, *MS4A6A*, and *GRN* are known risk genes for mental or behavioral disorders; *MS4A4A*, *SLC24A4*, *APOC2*, and *ZNF226* are known risk genes for AD biomarkers, lipoprotein related phenotypes (ApoB, LDL-C) and inflammatory-related phenotypes (CRP, inflammatory biomarkers).

Interestingly, the enriched phenotypes of ApoB, LDL-C, inflammatory biomarkers, and CRP have been reported to be associated with AD by previous studies. Elevated LDL-C levels have been implicated in AD development, as they can contribute to lipid-rich deposits in the brain, potentially leading to the formation of amyloid-beta plaques and neurofibrillary tangles (two major AD pathologies) [46]. In addition, as a major protein component of LDL-C, several studies have suggested a link between elevated ApoB levels and a higher risk of AD dementia [47, 48, 49, 50]. Chronic inflammation has been proposed as a significant risk factor in AD pathogenesis [51]. Increased levels of inflammatory biomarkers, such as cytokines and chemokines, have been observed in the brains of AD patients [52]. CRP is an acute-phase protein produced in response to inflammation. Elevated levels of CRP have been associated with an increased risk of AD dementia [53].

## Discussion

In this study, we applied BGW-TWAS to the reference GTEx V8 data of three tissues (prefrontal cortex, cortex, and whole blood) and employed ACAT-O to combine BGW-TWAS p-values across these three tissues to study AD dementia. We identified 37 significant genes with the prefrontal cortex, 55 significant genes with the cortex, and 51 significant genes with the whole blood reference transcriptomic data. Further, we obtained 93 TWAS risk genes of AD dementia with significant ACAT-O p-values. We also found that ∼31% of significant TWAS risk genes were primarily driven by *trans*-eQTL. Among these 93 identified TWAS risk genes, 9 were detected by TWAS using the TIGAR method [9, 11] with reference transcriptomic data of prefrontal cortex and the same GWAS summary data of AD dementia; 34 were identified as AD risk genes by previous TWAS or GWAS studies; and 50 are novel findings. As expected, most of the genes exhibiting significance due to *trans*-eQTL were not detected by previous studies which failed to account for *trans*-eQTL (except for *TRAPPC6A*, *DMPK* and *DACT3* on chromosome 19). Interestingly, previous studies reported AD-related biological functions for example novel TWAS risk genes *UGG1, BAG6, DHX16*, and *TUBB* [32, 33, 34, 35, 36, 37].

Through protein-protein interaction network analysis using STRING, we identified 5 network clusters containing both known and novel AD risk genes. Our findings are consistent with previous studies highlighting the critical involvement of *APOE* on chromosome 19 in AD [8, 27], as evidenced by its extensive connectivity with other known risk genes within the network clusters. The PPI networks also underline the important function of MHC class I protein binding genes (TWAS risk genes *MS4A4A* and *MS4A6A* identified in our study) related to AD dementia as these genes are interconnected with *APOE* [38]. Also, we identified several PPI clusters mainly comprising novel TWAS risk genes of AD dementia. For example, one such cluster included novel risk genes *ZBTB22* and *SLC39A7*, which were found associated with zinc finger generation and played a role in AD progression [44]. Another cluster featured known AD risk genes *MTCH2* and *NDUFS2*, which are related to the oxidative phosphorylation process of mitochondrial metabolism [17, 45]. Overall, a total of 19 novel genes are connected with known AD risk genes involved in the identified PPI clusters. Further research is still needed to understand the roles of these novel findings in the biological mechanism of AD.

We also identified 7 phenotypes whose known risk genes were enriched in our identified 93 TWAS risk genes, including family history of AD, mental or behavioral disorders, biomarkers for AD, ApoB, LDL-C, inflammatory biomarkers, and CRP. Especially, ApoB, LDL-C, inflammatory biomarkers, and CRP have been reported to be associated with AD by previous studies [46, 47, 48, 49, 50, 51, 52, 53], suggesting a complex interplay between genetic, transcriptomic, metabolic, and inflammatory risk factors in the pathogenesis of AD. Shared risk genes such as *APOE*, *APOC2*, and *MS4A4A* [54, 55, 56] between these phenotypes and AD have also been reported by previous studies. Further studies are still needed to elucidate the shared mechanisms underlying these phenotypes and AD.

Nevertheless, our study has several limitations. First, due to the computation burden of running BGW-TWAS tool, we only applied BGW-TWAS to reference GTEx V8 data of three tissues (prefrontal cortex, cortex, and whole blood). Other tissues such as hippocampus, muscle, and spinal cord, are also known to play crucial roles in the biological mechanisms of AD dementia [57, 58, 59]. Failing to consider all available tissues in GTEx V8 data may fail to capture a full spectrum of genetically regulated gene expression associated with AD dementia. We have been working on further improving the computation efficiency of the BGW-TWAS tool and applying it to all available tissues in GTEx V8.

Second, although the summary-level GWAS data of AD dementia were meta-analysis results of multiple studies with diverse populations, the GTEx reference transcriptomic data used in this study mainly consist of individuals of European descent. Since eQTL effect sizes might differ across populations due to factors such as population-specific linkage disequilibrium patterns and different allele frequencies, it would be worth exploring the results of using reference transcriptomic data and testing GWAS data of the same population [60].

Third, the BVSR model employed in the BGW-TWAS method inherently assumes a sparse model implying that only a small number of eQTL have true causal effects on gene expression. Although this assumption can be computationally advantageous, it may not always accurately represent the underlying genetic architecture of complex gene expression quantitative traits. In our future work, we will also investigate using other statistical models for genome-wide TWAS analysis.

## Conclusions

In conclusion, our study highlights the importance of considering both *cis*- and *trans*-eQTL in TWAS analysis as it can help identify significant risk genes that would have been missed by using only *cis*-eQTL. We identified several well-known AD risk genes as well as novel genes that have potential associations with AD dementia and are interconnected with known AD risk genes in the same PPI networks. As a genome-wide TWAS, our study is the first to utilize both *cis*- and *trans*- eQTLs of multiple tissues for AD risk gene identification. Our results provide further insights into the underlying biological mechanisms of AD dementia and could help with the development of potential therapeutic targets.

## Supporting information

Supplemental Table1 and legends for supplemental figures

## Data Availability

The RNAseq transcriptomic and WGS genetic data of GTEx V8 are publicly available from dbGAP (phs000424.v8.p2) and the GTEx portal website (https://gtexportal.org/home/). The summary-level GWAS data of AD dementia were generated by Wightman D.P. et. al. by meta-analysis excluding 23andMe samples, which are publicly available at https://ctg.cncr.nl/software/summary_statistics.

https://ctg.cncr.nl/software/summary_statistics

## List of Abbreviations

AD: Alzheimer’s disease
GWAS: Genome-wide association studies
TWAS: Transcriptome-wide association study
BGW-TWAS: Bayesian Genome-wide TWAS
eQTL: Expression quantitative trait loci
GTEx: Genotype-Tissue Expression
ACAT-O: Omnibus aggregated Cauchy association test
GReX: Genetically regulated gene expression
BVSR: Bayesian variable selection regression
EM-MCMC: Expectation-maximization Markov Chain Monte Carlo
PCP: Posterior causal probabilities
WGS: Whole genome sequencing
TPM: Transcripts Per Million
PPI: Protein-protein interaction
MS4: Membrane-spanning 4-domains
ApoB: Apolipoprotein b
LDL-C: Low-density lipoprotein cholesterol
CRP: C-reactive protein

## Declarations

### Ethics approval and consent to participate

All genomic data analyzed during the course of this study are either de-identified or summary statistics, which are considered as human data according to the guidelines of National Institute of Health.

### Consent for publication

Not applicable.

### Availability of data and materials

The RNAseq transcriptomic and WGS genetic data of GTEx V8 are publicly available from dbGAP (phs000424.v8.p2) and the GTEx portal website (https://gtexportal.org/home/). The summary-level GWAS data of AD dementia were generated by Wightman D.P. et. al. by meta-analysis excluding 23andMe samples, which are publicly available at https://ctg.cncr.nl/software/summary_statistics. The BGW-TWAS tool is available freely on GitHub (https://github.com/yanglab-emory/BGW-TWAS). The STRING tool is available online (https://string-db.org/). Summary data of our BGW-TWAS results will be shared with the public through SYNAPSE when this manuscript is accepted for publication.

### Authors’ contributions

S.G. analyzed the data, drafted and revised the manuscript. J.Y. designed the study and revised the manuscript.

## Acknowledgements

We would like to thank Dr. Michael P. Epstein from the Center for Computational and Quantitative Genetics, Department of Human Genetics, Emory University School of Medicine, for his helpful comments on this work.

## References

1. 2022 Alzheimer’s disease facts and figures. Alzheimers Dement. 2022;18(4):700–89.

2. Long JM, Holtzman DM. Alzheimer Disease: An Update on Pathobiology and Treatment Strategies. Cell. 2019;179(2):312–39.

3. Gatz M, Reynolds CA, Fratiglioni L, Johansson B, Mortimer JA, Berg S, et al. Role of genes and environments for explaining Alzheimer disease. Arch Gen Psychiatry. 2006;63(2):168–74.

4. Jansen IE, Savage JE, Watanabe K, Bryois J, Williams DM, Steinberg S, et al. Genome-wide meta-analysis identifies new loci and functional pathways influencing Alzheimer’s disease risk. Nat Genet. 2019;51(3):404–13.

5. Wightman DP, Jansen IE, Savage JE, Shadrin AA, Bahrami S, Holland D, et al. A genome-wide association study with 1,126,563 individuals identifies new risk loci for Alzheimer’s disease. Nature genetics. 2021;53(9):1276–82.

6. Gamazon ER, Wheeler HE, Shah KP, Mozaffari SV, Aquino-Michaels K, Carroll RJ, et al. A gene-based association method for mapping traits using reference transcriptome data. Nat Genet. 2015;47(9):1091–8.

7. Zhao BX, Shan Y, Yang Y, Yu ZL, Li TF, Wang XF, et al. Transcriptome-wide association analysis of brain structures yields insights into pleiotropy with complex neuropsychiatric traits. Nature Communications. 2021;12(1).

8. Sun Y, Zhu J, Zhou D, Canchi S, Wu C, Cox NJ, et al. A transcriptome-wide association study of Alzheimer’s disease using prediction models of relevant tissues identifies novel candidate susceptibility genes. Genome Med. 2021;13(1):141.

9. Parrish RL, Gibson GC, Epstein MP, Yang J. TIGAR-V2: Efficient TWAS tool with nonparametric Bayesian eQTL weights of 49 tissue types from GTEx V8. HGG Adv. 2022;3(1):100068.

10. Gusev A, Ko A, Shi H, Bhatia G, Chung W, Penninx BW, et al. Integrative approaches for large-scale transcriptome-wide association studies. Nat Genet. 2016;48(3):245–52.

11. Nagpal S, Meng X, Epstein MP, Tsoi LC, Patrick M, Gibson G, et al. TIGAR: An Improved Bayesian Tool for Transcriptomic Data Imputation Enhances Gene Mapping of Complex Traits. Am J Hum Genet. 2019;105(2):258–66.

12. Vosa U, Claringbould A, Westra HJ, Bonder MJ, Deelen P, Zeng B, et al. Large-scale cis- and trans-eQTL analyses identify thousands of genetic loci and polygenic scores that regulate blood gene expression. Nat Genet. 2021;53(9):1300–10.

13. Westra HJ, Peters MJ, Esko T, Yaghootkar H, Schurmann C, Kettunen J, et al. Systematic identification of trans eQTLs as putative drivers of known disease associations. Nat Genet. 2013;45(10):1238–43.

14. Luningham JM, Chen J, Tang S, De Jager PL, Bennett DA, Buchman AS, et al. Bayesian Genome-wide TWAS Method to Leverage both cis- and trans-eQTL Information through Summary Statistics. Am J Hum Genet. 2020;107(4):714–26.

15. Consortium GT. The GTEx Consortium atlas of genetic regulatory effects across human tissues. Science. 2020;369(6509):1318–30.

16. Serrano-Pozo A, Frosch MP, Masliah E, Hyman BT. Neuropathological alterations in Alzheimer disease. Cold Spring Harb Perspect Med. 2011;1(1):a006189.

17. Lunnon K, Keohane A, Pidsley R, Newhouse S, Riddoch-Contreras J, Thubron EB, et al. Mitochondrial genes are altered in blood early in Alzheimer’s disease. Neurobiol Aging. 2017;53:36–47.

18. Souza VC, Morais GS, Jr., Henriques AD, Machado-Silva W, Perez DIV, Brito CJ, et al. Whole-Blood Levels of MicroRNA-9 Are Decreased in Patients With Late-Onset Alzheimer Disease. Am J Alzheimers Dis Other Demen. 2020;35:1533317520911573.

19. Sullivan PF, Fan C, Perou CM. Evaluating the comparability of gene expression in blood and brain. Am J Med Genet B Neuropsychiatr Genet. 2006;141B(3):261–8.

20. Barbeira AN, Dickinson SP, Bonazzola R, Zheng J, Wheeler HE, Torres JM, et al. Exploring the phenotypic consequences of tissue specific gene expression variation inferred from GWAS summary statistics. Nat Commun. 2018;9(1):1825.

21. Liu Y, Chen S, Li Z, Morrison AC, Boerwinkle E, Lin X. ACAT: A Fast and Powerful p Value Combination Method for Rare-Variant Analysis in Sequencing Studies. Am J Hum Genet. 2019;104(3):410–21.

22. Szklarczyk D, Gable AL, Nastou KC, Lyon D, Kirsch R, Pyysalo S, et al. The STRING database in 2021: customizable protein-protein networks, and functional characterization of user-uploaded gene/measurement sets. Nucleic Acids Res. 2021;49(D1):D605–D12.

23. Yongtao Guan MS. Bayesian variable selection regression for genome-wide association studies and other large-scale problems. Ann Appl Stat. 2011;5(3):1780–815.

24. Yang J, Fritsche LG, Zhou X, Abecasis G, International Age-Related Macular Degeneration Genomics C. A Scalable Bayesian Method for Integrating Functional Information in Genome-wide Association Studies. Am J Hum Genet. 2017;101(3):404–16.

25. Svishcheva GR, Belonogova NM, Zorkoltseva IV, Kirichenko AV, Axenovich TI. Gene-based association tests using GWAS summary statistics. Bioinformatics. 2019;35(19):3701–8.

26. Hu T, Parrish RL, Buchman AS, Tasaki S, Bennett DA, Seyfried NT, et al. Omnibus proteome-wide association study (PWAS-O) identified 43 risk genes for Alzheimer’s disease dementia. medRxiv. 2022:2022.12.25.22283936.

27. Tang S, Buchman AS, De Jager PL, Bennett DA, Epstein MP, Yang J. Novel Variance-Component TWAS method for studying complex human diseases with applications to Alzheimer’s dementia. PLoS Genet. 2021;17(4):e1009482.

28. De Jager PL, Ma Y, McCabe C, Xu J, Vardarajan BN, Felsky D, et al. A multi-omic atlas of the human frontal cortex for aging and Alzheimer’s disease research. Sci Data. 2018;5:180142.

29. Gockley J, Montgomery KS, Poehlman WL, Wiley JC, Liu Y, Gerasimov E, et al. Multi-tissue neocortical transcriptome-wide association study implicates 8 genes across 6 genomic loci in Alzheimer’s disease. Genome Med. 2021;13(1):76.

30. Liu N, Xu J, Liu H, Zhang S, Li M, Zhou Y, et al. Hippocampal transcriptome-wide association study and neurobiological pathway analysis for Alzheimer’s disease. PLoS Genet. 2021;17(2):e1009363.

31. Rao S, Ghani M, Guo Z, Deming Y, Wang K, Sims R, et al. An APOE-independent cis-eSNP on chromosome 19q13.32 influences tau levels and late-onset Alzheimer’s disease risk. Neurobiol Aging. 2018;66:178 e1–e8.

32. Suzuki M, Tezuka K, Handa T, Sato R, Takeuchi H, Takao M, et al. Upregulation of ribosome complexes at the blood-brain barrier in Alzheimer’s disease patients. J Cereb Blood Flow Metab. 2022;42(11):2134–50.

33. Kasu YAT, Arva A, Johnson J, Sajan C, Manzano J, Hennes A, et al. BAG6 prevents the aggregation of neurodegeneration-associated fragments of TDP43. iScience. 2022;25(5):104273.

34. al VFe. An enrichment of rare variants and the lysosomal pathways are important contributors to early onset Alzheimer disease. Alzheimer’s & Dementia. 2021;17(S3).

35. Xiong W, Cai J, Li R, Wen C, Tan H, On Behalf Of The Alzheimer’s Disease Neuroimaging Initiative Adni D. Rare Variant Analysis and Molecular Dynamics Simulation in Alzheimer’s Disease Identifies Exonic Variants in FLG. Genes (Basel). 2022;13(5).

36. Duke CG, Kennedy AJ, Gavin CF, Day JJ, Sweatt JD. Experience-dependent epigenomic reorganization in the hippocampus. Learn Mem. 2017;24(7):278–88.

37. Puig B, Ferrer I, Luduena RF, Avila J. BetaII-tubulin and phospho-tau aggregates in Alzheimer’s disease and Pick’s disease. J Alzheimers Dis. 2005;7(3):213–20; discussion 55-62.

38. Antunez C, Boada M, Gonzalez-Perez A, Gayan J, Ramirez-Lorca R, Marin J, et al. The membrane-spanning 4-domains, subfamily A (MS4A) gene cluster contains a common variant associated with Alzheimer’s disease. Genome Med. 2011;3(5):33.

39. Lazarczyk MJ, Kemmler JE, Eyford BA, Short JA, Varghese M, Sowa A, et al. Major Histocompatibility Complex class I proteins are critical for maintaining neuronal structural complexity in the aging brain. Sci Rep. 2016;6:26199.

40. Zalocusky KA, Najm R, Taubes AL, Hao Y, Yoon SY, Koutsodendris N, et al. Neuronal ApoE upregulates MHC-I expression to drive selective neurodegeneration in Alzheimer’s disease. Nat Neurosci. 2021;24(6):786–98.

41. Tan MS, Yang YX, Xu W, Wang HF, Tan L, Zuo CT, et al. Associations of Alzheimer’s disease risk variants with gene expression, amyloidosis, tauopathy, and neurodegeneration. Alzheimers Res Ther. 2021;13(1):15.

42. Nho K, Kim S, Horgusluoglu E, Risacher SL, Shen L, Kim D, et al. Association analysis of rare variants near the APOE region with CSF and neuroimaging biomarkers of Alzheimer’s disease. BMC Med Genomics. 2017;10(Suppl 1):29.

43. Hamilton G, Harris SE, Davies G, Liewald DC, Tenesa A, Starr JM, et al. Alzheimer’s disease genes are associated with measures of cognitive ageing in the lothian birth cohorts of 1921 and 1936. Int J Alzheimers Dis. 2011;2011:505984.

44. Bu S, Lv Y, Liu Y, Qiao S, Wang H. Zinc Finger Proteins in Neuro-Related Diseases Progression. Front Neurosci. 2021;15:760567.

45. Ruggiero A, Aloni E, Korkotian E, Zaltsman Y, Oni-Biton E, Kuperman Y, et al. Loss of forebrain MTCH2 decreases mitochondria motility and calcium handling and impairs hippocampal-dependent cognitive functions. Sci Rep. 2017;7:44401.

46. Di Paolo G, Kim TW. Linking lipids to Alzheimer’s disease: cholesterol and beyond. Nat Rev Neurosci. 2011;12(5):284–96.

47. Wingo TS, Cutler DJ, Wingo AP, Le NA, Rabinovici GD, Miller BL, et al. Association of Early-Onset Alzheimer Disease With Elevated Low-Density Lipoprotein Cholesterol Levels and Rare Genetic Coding Variants of APOB. JAMA Neurol. 2019;76(7):809–17.

48. Hu H, Tan L, Bi YL, Xu W, Tan L, Shen XN, et al. Association of serum Apolipoprotein B with cerebrospinal fluid biomarkers of Alzheimer’s pathology. Ann Clin Transl Neurol. 2020;7(10):1766–78.

49. Button EB, Gilmour M, Cheema HK, Martin EM, Agbay A, Robert J, et al. Vasoprotective Functions of High-Density Lipoproteins Relevant to Alzheimer’s Disease Are Partially Conserved in Apolipoprotein B-Depleted Plasma. Int J Mol Sci. 2019;20(3).

50. Picard C, Nilsson N, Labonte A, Auld D, Rosa-Neto P, Alzheimer’s Disease Neuroimaging I, et al. Apolipoprotein B is a novel marker for early tau pathology in Alzheimer’s disease. Alzheimers Dement. 2022;18(5):875–87.

51. Heneka MT, Carson MJ, El Khoury J, Landreth GE, Brosseron F, Feinstein DL, et al. Neuroinflammation in Alzheimer’s disease. Lancet Neurol. 2015;14(4):388–405.

52. Holmes C, Cunningham C, Zotova E, Woolford J, Dean C, Kerr S, et al. Systemic inflammation and disease progression in Alzheimer disease. Neurology. 2009;73(10):768–74.

53. Locascio JJ, Fukumoto H, Yap L, Bottiglieri T, Growdon JH, Hyman BT, et al. Plasma amyloid beta-protein and C-reactive protein in relation to the rate of progression of Alzheimer disease. Arch Neurol. 2008;65(6):776–85.

54. Serrano-Pozo A, Das S, Hyman BT. APOE and Alzheimer’s disease: advances in genetics, pathophysiology, and therapeutic approaches. Lancet Neurol. 2021;20(1):68–80.

55. Elliott DA, Weickert CS, Garner B. Apolipoproteins in the brain: implications for neurological and psychiatric disorders. Clin Lipidol. 2010;51(4):555–73.

56. You SF, Brase L, Filipello F, Iyer AK, Del-Aguila J, He J, et al. MS4A4A modifies the risk of Alzheimer disease by regulating lipid metabolism and immune response in a unique microglia state. medRxiv. 2023.

57. Braak H, Braak E. Neuropathological stageing of Alzheimer-related changes. Acta Neuropathol. 1991;82(4):239–59.

58. Manczak M, Calkins MJ, Reddy PH. Impaired mitochondrial dynamics and abnormal interaction of amyloid beta with mitochondrial protein Drp1 in neurons from patients with Alzheimer’s disease: implications for neuronal damage. Hum Mol Genet. 2011;20(13):2495–509.

59. Pechlivanidou M, Kousiappa I, Angeli S, Sargiannidou I, Koupparis AM, Papacostas SS, et al. Glial Gap Junction Pathology in the Spinal Cord of the 5xFAD Mouse Model of Early-Onset Alzheimer’s Disease. Int J Mol Sci. 2022;23(24).

60. Bhattacharya A, Hirbo JB, Zhou D, Zhou W, Zheng J, Kanai M, et al. Best practices for multi-ancestry, meta-analytic transcriptome-wide association studies: Lessons from the Global Biobank Meta-analysis Initiative. Cell Genom. 2022;2(10).

