## Supplemental Table1 and legends for supplemental figures for "Bayesian genome-wide TWAS with reference transcriptomic data of brain and blood tissues identified 93 risk genes for Alzheimer’s disease dementia"

| Remaining Significant TWAS Risk Genes | | |
|---|---|---|
| Gene | CHR | p-value |
| NDUFS2[c] | 1 | 6.12E-07 |
| FCER1G[c] | 1 | 1.43E-07 |
| WDR33[c] | 2 | 1.52E-09 |
| UGGT1[b] | 2 | 5.66E-10 |
| TNIP1[c] | 5 | 2.19E-06 |
| TIMD4[b] | 5 | 8.52E-07 |
| CLIC1[b] | 6 | 2.33E-06 |
| HSD17B8[b] | 6 | 1.51E-06 |
| CFB[c] | 6 | 8.45E-07 |
| SLC39A7[a] | 6 | 6.05E-07 |
| ZBTB22[b] | 6 | 3.73E-08 |
| VPS52[b] | 6 | 1.27E-08 |
| GAL3ST4[c] | 7 | 9.79E-07 |
| EPHB4[c] | 7 | 6.67E-07 |
| PVRIG[b] | 7 | 1.49E-07 |
| AZGP1[b] | 7 | 2.84E-11 |
| OSBP[c] | 11 | 8.09E-08 |
| AP000442.1[b] | 11 | 7.53E-08 |
| CREBZF[b] | 11 | 6.72E-09 |
| TMEM132A[b] | 11 | 1.11E-11 |
| STYX[c] | 14 | 1.88E-07 |
| SLC24A4[c] | 14 | 6.08E-08 |
| ERCC1[c] | 19 | 1.76E-06 |
| AC092301.3[a] | 19 | 1.71E-06 |
| BCL3[c] | 19 | 1.40E-06 |
| CD33[c] | 19 | 9.86E-07 |
| ERCC2[c] | 19 | 5.09E-07 |
| AC092066.1[b] | 19 | 6.74E-09 |
| AC084219.4[c] | 19 | 4.71E-09 |
| ZNF225[b] | 19 | 1.83E-09 |
| ZNF180[a] | 19 | 8.10E-10 |
| ZNF221[b] | 19 | 1.39E-10 |
| ZNF296[b] | 19 | 5.69E-12 |
| FAM209B[c] | 20 | 8.60E-07 |

a: Genes significant for prefrontal cortex tissue
b: Genes significant for cortex tissue
c: Genes significant for whole blood tissue

**Supplemental Table 1: Remaining TWAS risk genes of AD dementia with significant ACAT-O p-values.**

### Supplemental Figure Legends

**Supplemental Figure 1. Venn diagram plot of the number of significant TWAS risk genes by BGW-TWAS with three tissues.** Purple denotes the number of genes identified by BGW-TWAS in prefrontal cortex tissue, green denotes the number of genes identified by BGW-TWAS in cortex tissue, and green denotes the number of genes identified by BGW-TWAS in whole blood tissue.

**Supplemental Figure 2: Manhattan plot of BGW-TWAS p-values of prefrontal cortex for studying AD dementia.** The horizontal dashed line represents the Bonferroni corrected significance threshold of the p-value, given by dividing 0.05 by the total number of test genes. Red dots represent genes that were significant in prefrontal cortex.

**Supplemental Figure 3: Manhattan plot of BGW-TWAS p-values of cortex for studying AD dementia.** The horizontal dashed line represents the Bonferroni corrected significance threshold of the p-value, given by dividing 0.05 by the total number of test genes. Red dots represent genes that were significant in cortex.

**Supplemental Figure 4: Manhattan plot of BGW-TWAS p-values of whole blood for studying AD dementia.** The horizontal dashed line represents the Bonferroni corrected significance threshold of the p-value, given by dividing 0.05 by the total number of test genes. Red dots represent genes that were significant in whole blood.

**Supplemental Figure 5. Proportions of *trans*-eQTL with non-zero weights of significant genes in three tissues.** A: prefrontal cortex tissue; B: cortex tissue; C: whole blood tissue. For each bar graph, the x-axis represents the proportion of *trans*-eQTL with non-zero weights, ranging from 0 to 1; the y-axis represents all the genes with significant BGW-TWAS p-values of the respective tissue, sorted by *trans*-eQTL proportions.

**Supplemental Figure 6: Venn diagram plot of the number of significant genes detected by ACAT-O using both *cis*-eQTL and *trans*-eQTL, and the number of significant genes detected by ACAT-O using only *cis*-eQTL.** Purple denotes using both *cis*- and *trans*- eQTL, and yellow denotes using only *cis*-eQTL.
