## Supplementary figures and images for "Bayesian genome-wide TWAS with reference transcriptomic data of brain and blood tissues identified 93 risk genes for Alzheimer’s disease dementia"

### Supplemental Figure 1

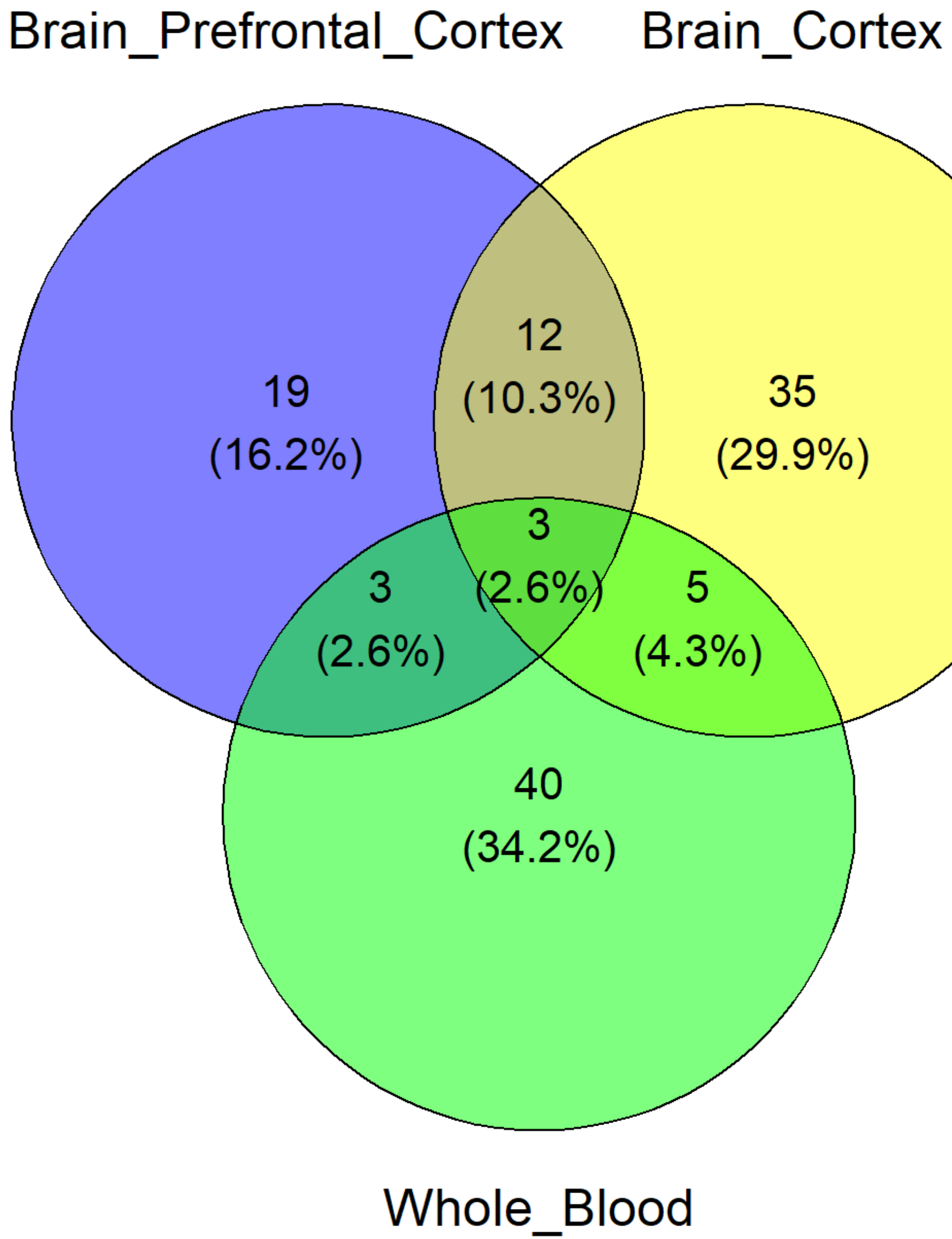

### Supplemental Figure 2

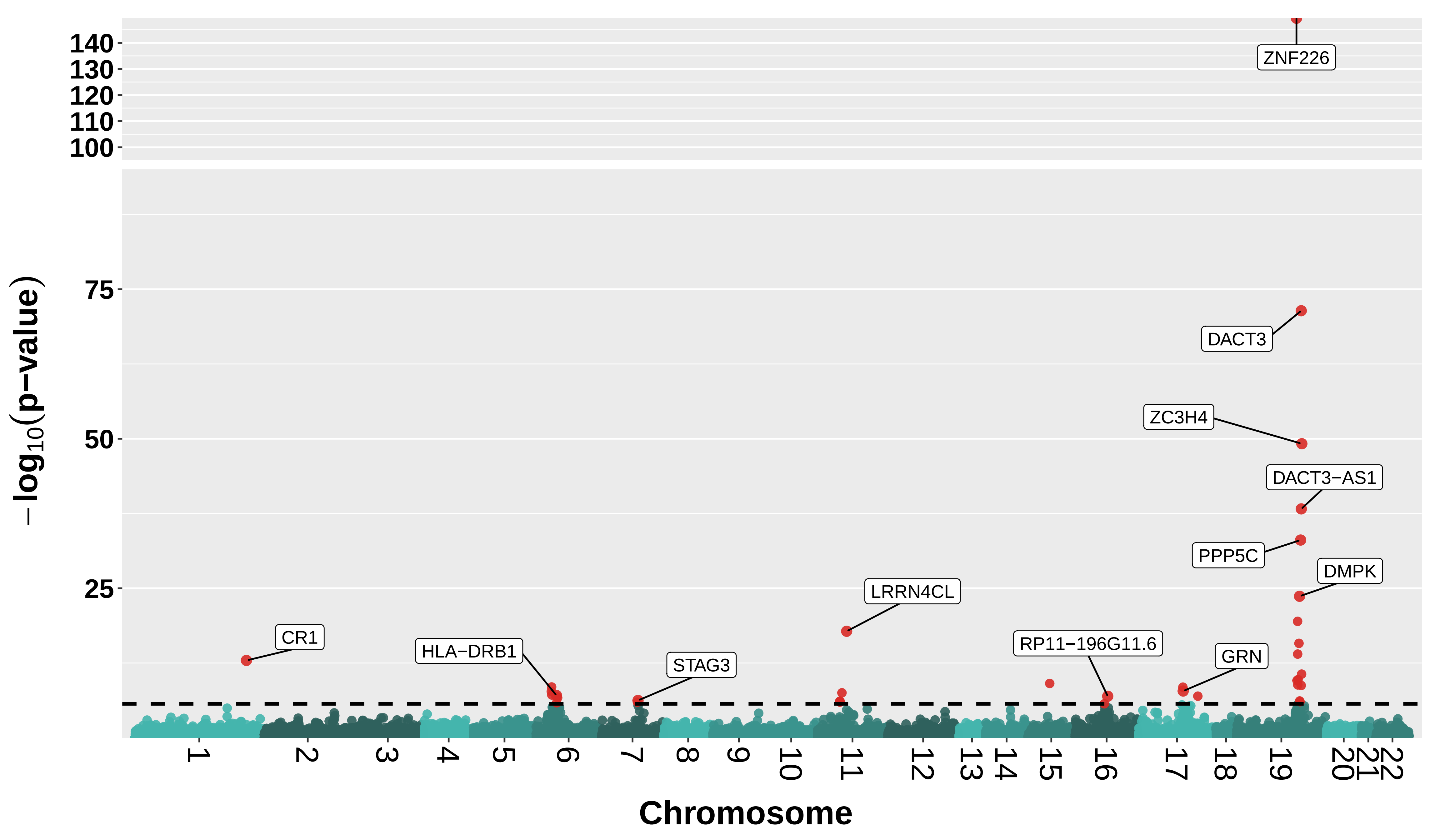

### Supplemental Figure 3

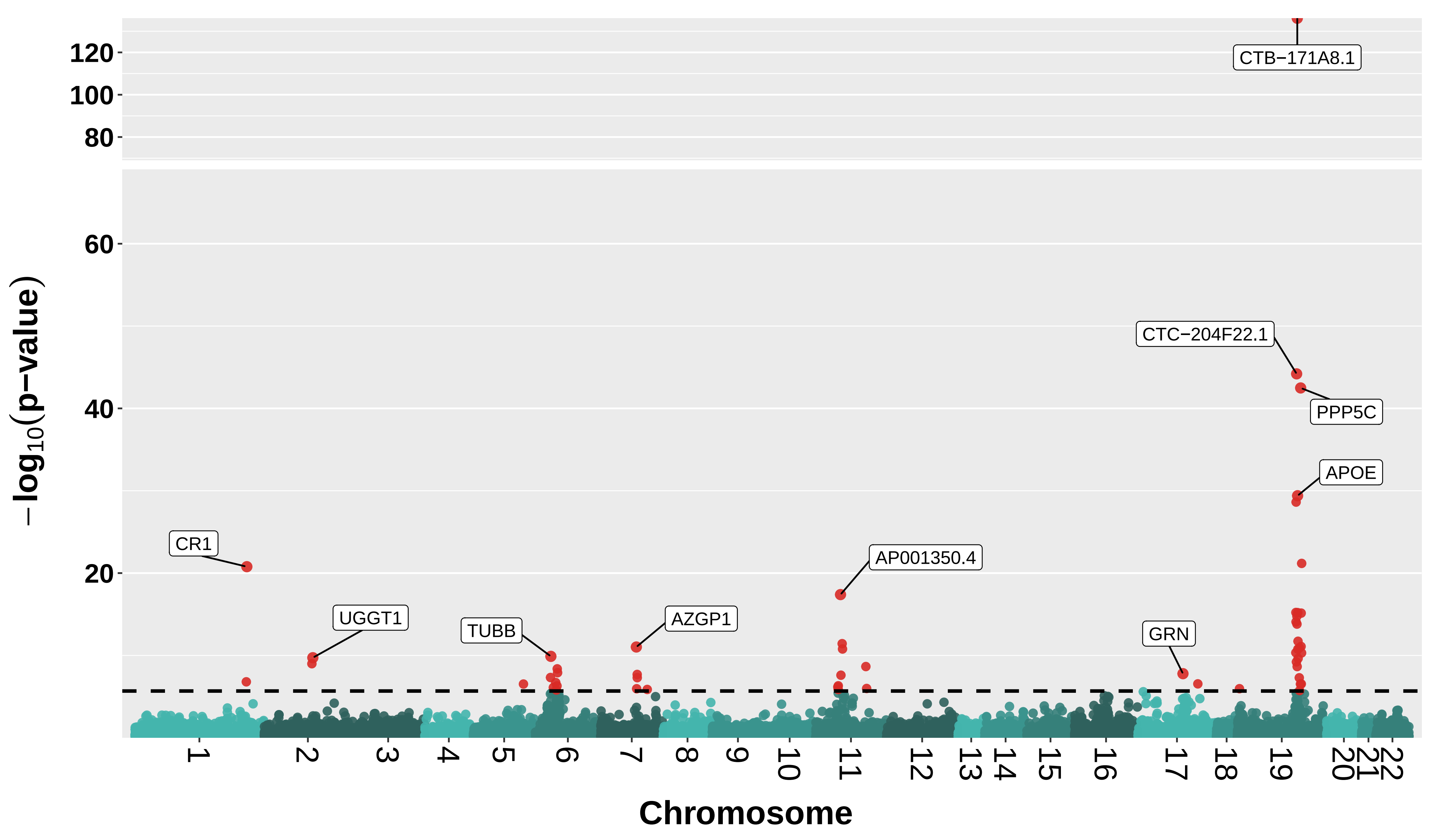

### Supplemental Figure 4

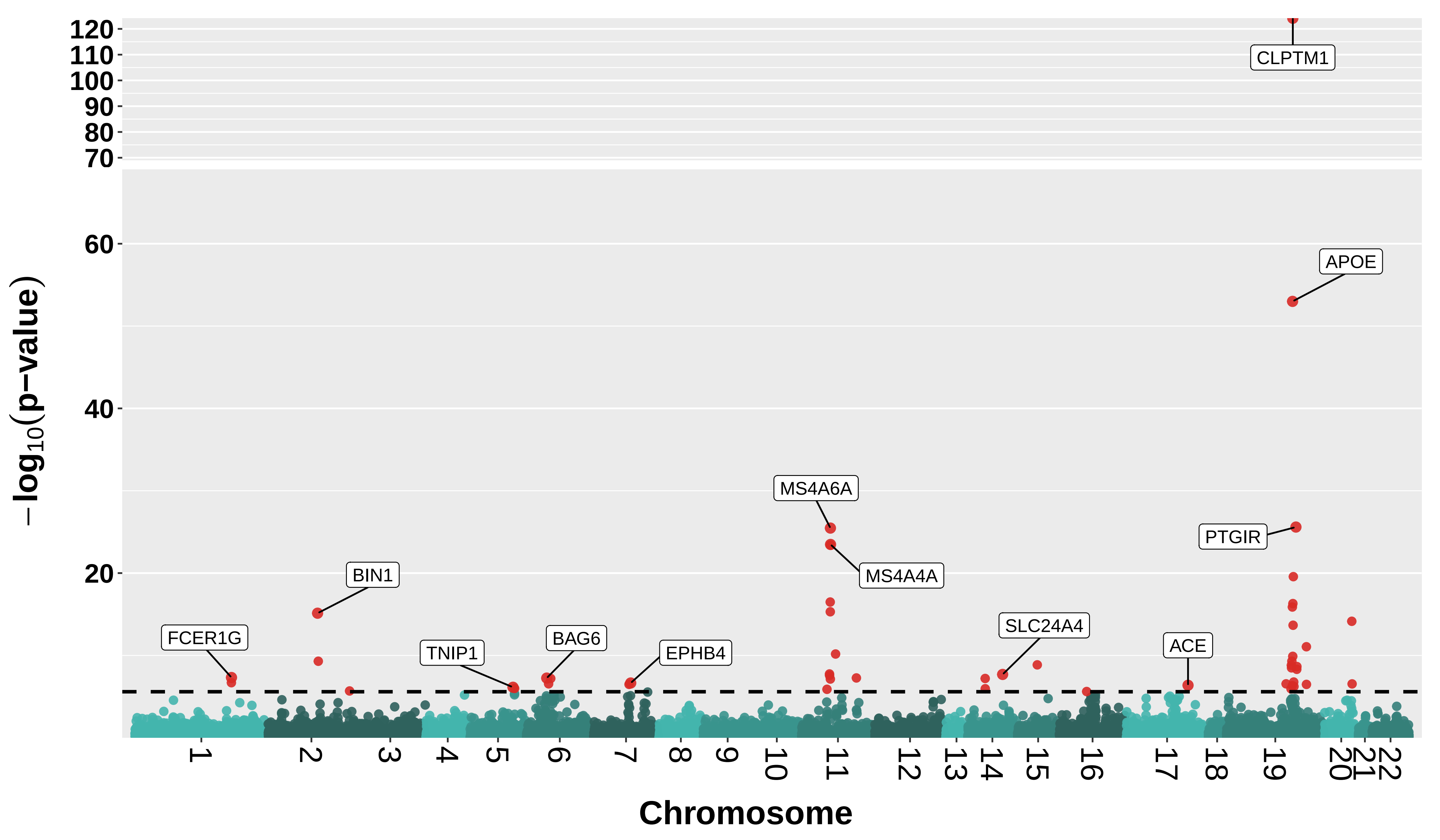

### Supplemental Figure 5

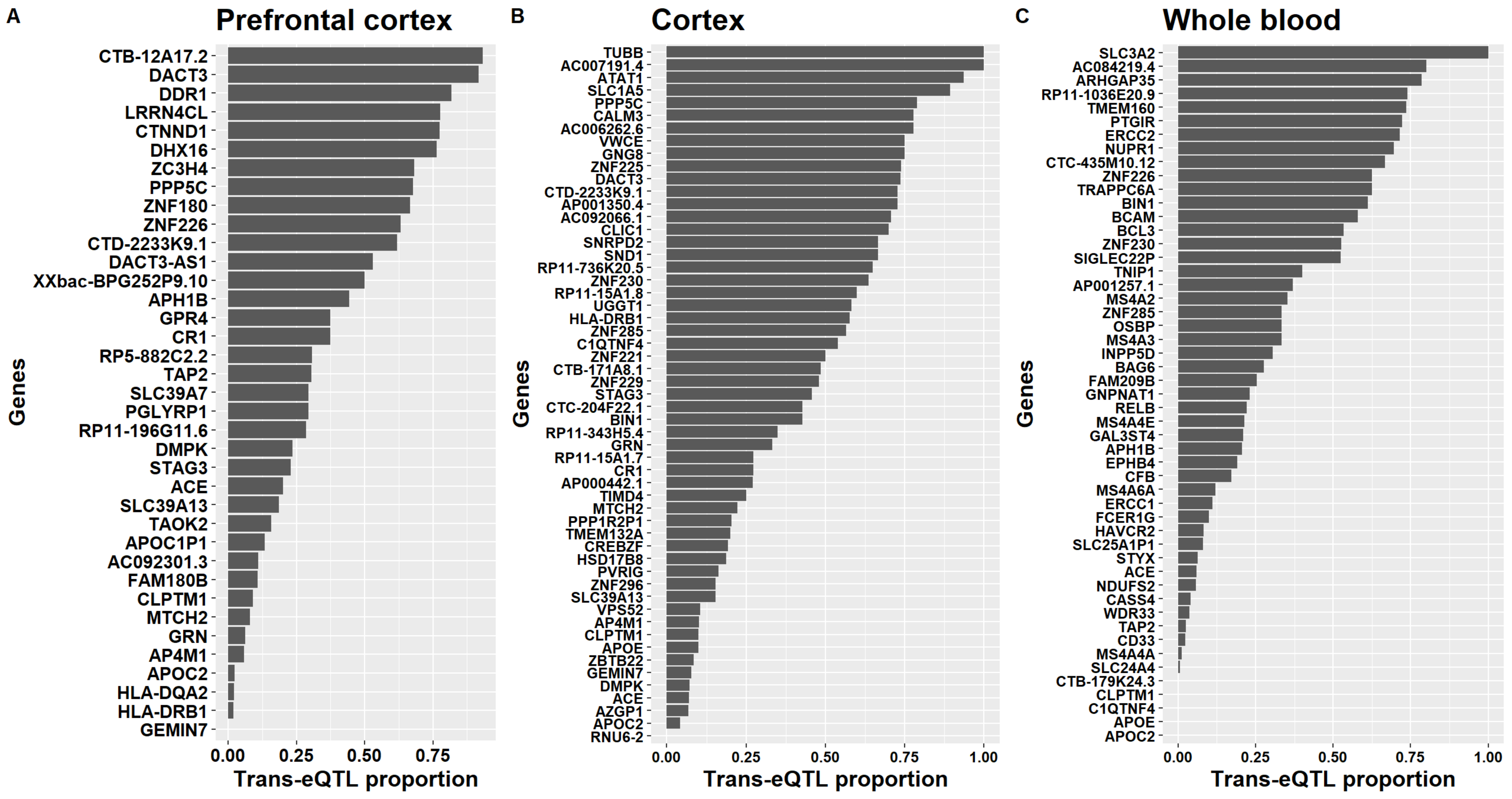

### Supplemental Figure 6

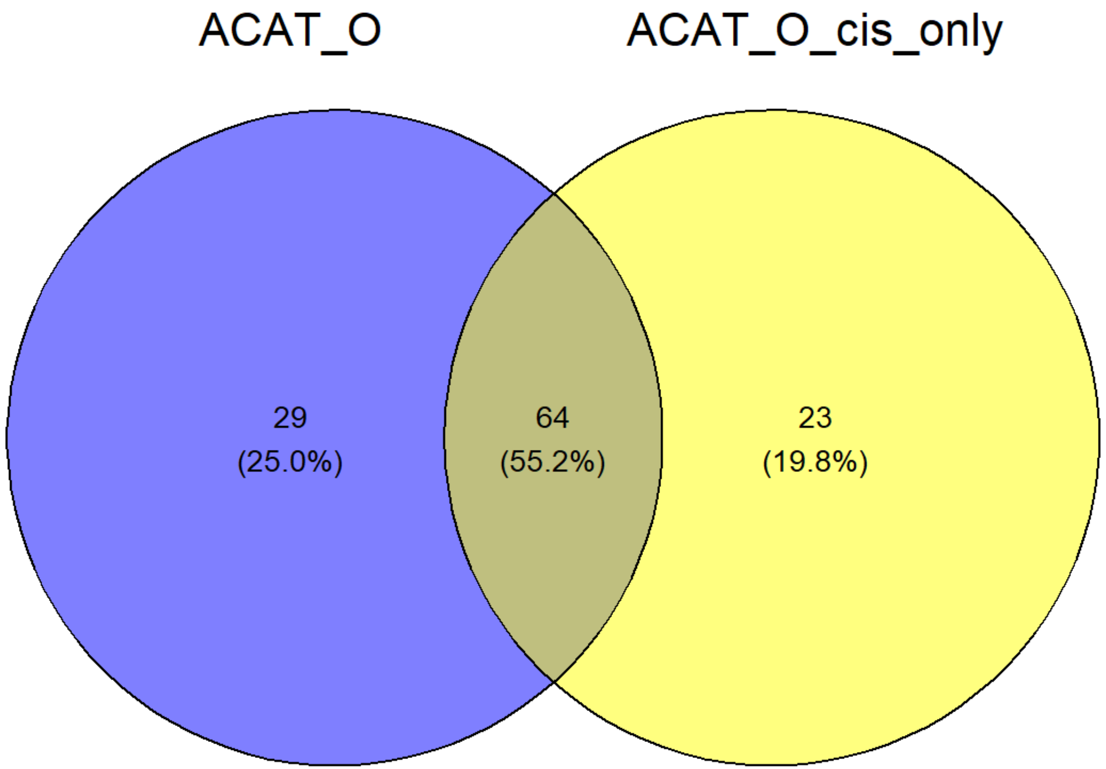
